# Within-person Relationships of Sleep Duration with Next-Day Stress and Affect in the Daily Life of Adults with Type-1 Diabetes

**DOI:** 10.1101/2023.01.22.23284883

**Authors:** Haomiao Jin, Jeffrey S Gonzalez, Elizabeth Pyatak, Stefan Schneider, Claire J Hoogendoorn, Raymond Hernandez, Pey-Jiuan Lee, Donna Spruijt-Metz

**Affiliations:** School of Health Sciences, University of Surrey, Guildford, UK; Division of Endocrinology, Department of Medicine, Albert Einstein College of Medicine, Bronx, NY, US; Ferkauf Graduate School of Psychology, Yeshiva University, Bronx, CA, US; Chan Division of Occupational Science and Occupational Therapy, University of Southern California, Los Angeles, CA, US; Center for Economic and Social Research, University of Southern California, Los Angeles, CA, US

**Keywords:** Type 1 diabetes, Sleep, Stress, Affect, Emotions, Longitudinal Studies

## Abstract

**Objective:** The objective of this study is to examine the within-person relationships between sleep duration and next-day stress and affect in the daily life of individuals with T1D.

**Methods:** Study participants were recruited in the Function and Emotion in Everyday Life with Type 1 Diabetes (FEEL-T1D) study. Sleep duration was derived by synthesizing objective (actigraphy) and self-report measures. General and diabetes-specific stress and positive and negative affect were measured using ecological momentary assessment. Multilevel regression was used to examine the within-person relationships between sleep duration and next-day stress and affect. Cross-level interactions were used to explore whether gender and baseline depression and anxiety moderated these within-person relationships.

**Results:** Adults with T1D (n=166) completed measurements for 14 days. The average age was 40.99 years, and 91 participants (54.82%) were female. The average sleep duration was 7.29 hours (SD=1.18 hours). Longer sleep was significantly associated with lower general stress (p<0.001) but not diabetes-specific stress (p=0.18) on the next day. There were significant within-person associations of longer sleep with lower levels on next-day negative affect (overall, p=0.002, disappoint, p=0.05; sad, p=0.05; tense, p<0.001; upset, p=0.008; anxious, p=0.04). There were no significant associations with positive affect. Examination of the interaction effects did not reveal significant differential relationships for men and women and for individuals with and without depression or anxiety at baseline.

**Conclusion:** Findings from this study suggest optimizing sleep duration as an important interventional target for better managing general stress and improving daily emotional wellbeing of individuals with T1D.

## 1. Introduction

Type 1 diabetes (T1D) is a common chronic disease affecting more than eight million individuals worldwide [1]. Individuals with T1D must cope with a wide range of challenges not only specific to the disease but also related to many other areas of their daily lives, leading to heightened stress and emotional burden [2–4]. Recommendations from the American Diabetes Association suggest the integration of psychosocial care, including the management of diabetes distress and emotional burden, with collaborative, patient-centered medical care to all individuals with diabetes [5].

Sleep is a necessary and crucial human function; nevertheless, sleep problems like insufficient sleep are common in present society [6,7]. Increasing research has shown that sleep duration is negatively related to stress and affective functioning in healthy adults [8–11]. Adults with T1D report more sleep disturbances than people without T1D [12]. Diabetes pathophysiology such as hypoglycemia, hyperglycemia and increased glucose variability have been suggested to negatively impact sleep [13–15].

Although the presence of T1D has a negative impact on stress, affect, and sleep, there has been little research on the within-person relationships between sleep and everyday stress and affect in the context of T1D. In a recent study, higher sleep quality was shown to be associated with fewer next-day general and T1D-specific stress [16]; no study has specifically examined the impact of sleep duration on stress and positive or negative affect in the daily life of individuals with T1D. To fill in this knowledge gap, this paper examined the within-person relationships between sleep duration and next-day stress and positive and negative affect using intensive longitudinal data collected from the daily life of individuals with T1D.

Insufficient sleep may influence diabetes distress, general stress, positive affect (PA), and negative affect (NA) through behavioral and physiological pathways. One pathway is that insufficient sleep can incur additional metabolic cost and energy needs, resulting in increased food intake and energy conservation behaviors [17,18]. This may create additional self-management needs for individuals with T1D and could therefore result in higher diabetes distress. Furthermore, sleep loss can influence the hypothalamic–pituitary–adrenal (HPA) axis, leading to increased stress hormone levels [8,19]. Accordingly, we hypothesized that longer sleep duration would show negative within-person association with next-day general and diabetes-specific stress in individuals with T1D.

As for the relationship between sleep duration and affect, we hypothesized that longer sleep duration would show positive association with PA and negative association with NA on the next day. The cognitive-energy model suggests that sleep loss can negatively impact cognitive-energy resources that are required for coping with goal-obstructing events or for capitalizing on new opportunities offered by goal-enhancing events [20]. When having enough cognitive-energy resources to reach a goal is anticipated, PA is promoted; when a lack of resources is perceived, NA is increased. Existing research based on healthy adults suggested that while sleep is associated with both PA and NA, the association with PA is stronger than the association with NA [21–24].

In addition to testing the main hypotheses, we also conducted exploratory analyses for the differential relationships of sleep duration with next-day stress and affect based on gender and depression and anxiety status at baseline. A combination of environmental, social, and cultural differences cause gender differences in sleep behavior and sleep disorders [25]. Prevalence of sleep problems is higher among women than men with T1D [26], which exposes more women with T1D to the negative impacts of poor sleep. Nevertheless, whether sleep would have differential relationships with health-related outcomes like stress and affect in men and women with T1D remains unclear. Individuals with T1D are at increased risk for depression and anxiety [27,28]. Similar to the gender effect, depression and anxiety are also associated with increased sleep problems, [29,30]. In view of that, exploratory analysis was carried out to examine the potentially differential relationships based on gender and depression and anxiety status at baseline.

## 2. Methods

### 2.1. Study sample and data collection

The study sample included participants of the Function and Emotion in Everyday Life with Type 1 Diabetes (FEEL-T1D) study, a multiyear research project funded by the US National Institutes of Health (1R01DK121298). Participants were recruited between June 2020 and February 2022 from two large health systems in Los Angeles, California and the Bronx, New York via direct solicitations (phone, mail, email) from research coordinators. Participants were English-or Spanish-speaking adults 18 years or older, who had type 1 diabetes for at least one year, who were on a stable treatment regimen for diabetes and comorbid medical conditions, and who had sufficient visual acuity, cognition, and fine motor abilities to carry out the study protocol.

After enrollment, participants completed a series of demographic, clinical and psychosocial questionnaires and were subsequently mailed study materials. These included a wrist-worn wGT3X-BT accelerometer (Actigraph), blinded CGM (Abbott FreeStyle LibrePro Flash Glucose Monitoring System), and smartphone (Xiaomi Mi A1) preloaded with an ecological momentary assessment (EMA) survey application (Ilumivu) and mobile cognitive tasks [31]. After applying the study CGM and being trained in using the study devices, participants completed 14 days of intensive longitudinal data collection during which they completed EMA surveys and cognitive tasks through the day at 3-hr intervals. This led to 5-6 times of assessments per day depending on individual’s wake to sleep times. The full protocol for the FEEL-T1D study was previously published [32]. The study protocol was approved by the University of Southern California and Albert Einstein College of Medicine Institutional Review Boards. All participants provided informed consent prior to starting study procedures.

### 2.2. Measures

#### Sleep duration

Sleep and wake times were derived from two sources: (a) patient self-report data collected at the first (i.e., morning) EMA prompt of each day; (b) accelerometry data processed with ActiLife software version 6.13.4, using Tudor-Locke auto sleep period detection calculated by the Cole-Kripke algorithm [33]. As each of these sources can carry measurement error, we applied an algorithm to combine information from both sources [34]. The algorithm uses the empirically determined probability distribution of sleep and wake times for each individual to create a weighted average of self-reported and accelerometry-derived data for each individual and day, giving more weight to the empirically more likely value [34].

#### General and diabetes-specific stress

General stress level was measured by an EMA question that asked “how stressed are you right now?”. Diabetes-specific stress was measured by an EMA question that asked “how stressed do you feel about your diabetes or diabetes management right now?”. Responses were on a slider scale from 0 indicating “not at all stressed” to 100 indicating “extremely stressed”.

#### Affect

Affect was measured by EMA surveys with nine affect items derived from the Stress and Working Memory Study [35]. These items included four PA items (i.e., happy, content, enthusiastic, and excited) and five NA items (i.e., disappointed, sad, tense, upset, and anxious). An example question was “right now, how sad do you feel?”. The responses were scored from 0 to 100 with 0 indicating “not at all” and 100 indicating “extremely”. We included individual items as outcomes and also computed overall PA and NA measures as the average scores of the positive and negative affect items respectively.

#### Blood glucose

Blood glucose data were obtained from the Abbott Diabetes Scientific Research Group, who processed the LibrePro CGM sensors using the Libre2 algorithm. Mean and standard deviations were calculated for the time asleep as defined above and for the three-hour intervals between EMA assessments during daytime.

#### Other measures

Self-identified race, ethnicity, gender, marital status, education level, and annual income were collected via a study-specific demographic questionnaire administered at baseline. Depression at baseline was measured with the 8-item Patient Health Questionnaire; patients were categorized into subgroups using an established cutoff score of ≥10 [36]. Anxiety at baseline was measured with the 7-item Generalized Anxiety Disorder scale score; subgroups were defined using a ≥10 cutoff [37]. Pain was measured by an EMA question asking “at this moment, how much bodily pain do you feel?”. The response value was from 0 indicating “no pain” to 100 indicating “extreme pain”.

### 2.3. Statistical analysis

Participant characteristics including basic demographics, depression and anxiety at study baseline were summarized using descriptive statistics. Sleep duration was normalized using within-person z-scores [11], which was used as predictor of the measures of stress and affect on the next day to examine their relationships. A three-level multilevel regression model was used that clustered EMA responses (Level 1) of the outcome variable (stress or affect) by study days (Level 2) and participants (Level 3). Given that sleep duration was a day-level (Level 2) predictor variable, we could alternatively have computed the average of the 5-6 EMA responses per day and used the computed averages as outcome variables in a simpler 2-level model. However, a 3-Level model appropriately accounts for measurement error in the EMA responses (where 5-6 moments were sampled from all moments of the day) and accommodates unbalanced data due to missed EMA prompts.

Covariates in the models included day of week as a time-varying categorical covariate, within-person normalized pain levels, and mean and standard deviation of blood glucose of the time asleep and three-hour intervals. Because the aim of our analysis was to examine within-person relationships, there was no need to control for time-invariant (i.e., between-person) covariates. However, we examined the cross-level interaction effects of sleep duration with gender and depression and anxiety at baseline to understand whether differential relationships existed for men and women and for T1D patients with and without mental health conditions. Random effects included the intercept and the slope of sleep duration. As longer durations of sleep can also have negative effects on well-being [38], we examined both linear and curvilinear relationships using orthogonal polynomials. We reported results from the linear models since no higher-order terms showed significant effects. All statistical analysis in this paper was carried out using the R package “lme4” [39].

## 3. Results

The study participants included 166 individuals with T1D. As shown in Table 1, the average age was 39.55 years, and 91 participants (54.82%) were female. At study baseline, 24 participants (14.46%) had depression as assessed by PHQ-8 score ≥ 10, and 13 participants (7.83%) had anxiety as assessed by GAD-7 score ≥ 10. The average sleep duration was 7.29 hours (SD=1.18 hours).

**Table 1.**
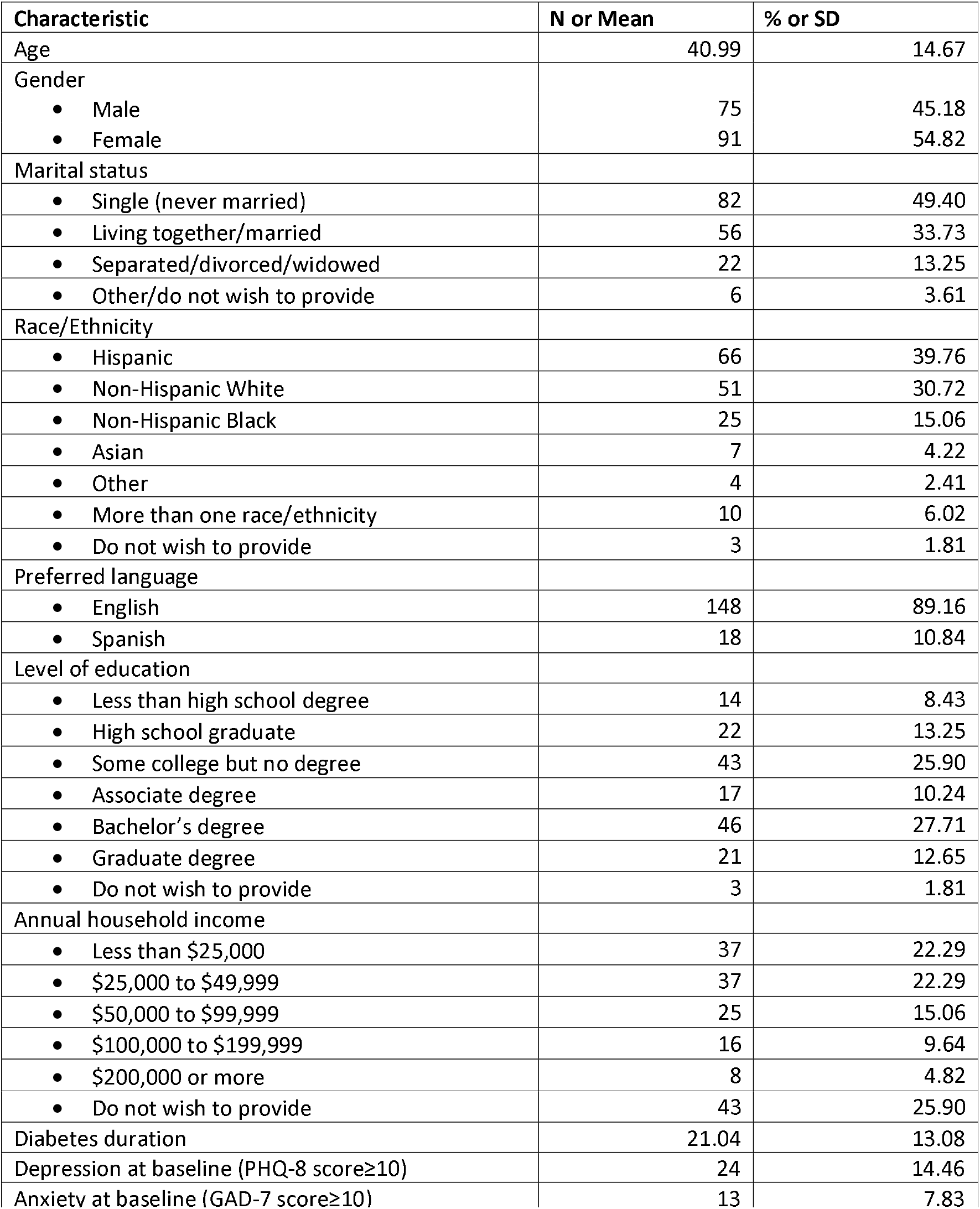
Baseline characteristics of Participants in the Function and Emotion in Everyday Life with Type 1 Diabetes (FEEL-T1D) Study (N=166)

The estimates of within-person relationships between sleep duration and next-day stress, PA, NA are shown in Table 2. Longer sleep was significantly associated with lower general stress (coefficient: -1.31, 95% confidence interval [CI]: -1.80 ∼ -0.83, p < 0.001) but not diabetes-specific stress (coefficient: -0.40, 95% CI: -0.98 ∼ 0.19, p=0.18). Longer sleep was significantly associated with lower next-day NA (overall NA, coefficient: -0.76, 95% CI: -1.25 ∼ -0.28, p = 0.002; disappoint, coefficient: -0.62, 95% CI: -1.22 ∼ 0.00, p = 0.05; sad, coefficient: -0.67, 95% CI: -1.35 ∼ 0.00, p = 0.05; tense, coefficient: -1.14, 95% CI: -1.79 ∼ -0.50, p < 0.001; upset, coefficient: -0.79, 95% CI: -1.36 ∼ - 0.21, p = 0.008; anxious, coefficient: -0.69, 95% CI: -1.34, -0.04, p = 0.04). There were no significant associations with any PA variables. The estimates for covariates in the models for general stress and overall NA are shown in Table 3 (estimates for covariates in all models are shown in the Appendix Table). Among the covariates, greater levels of pain were significantly associated with more NA and higher general stress. The general stress were lower during weekends compared with Monday.

**Table 2.**
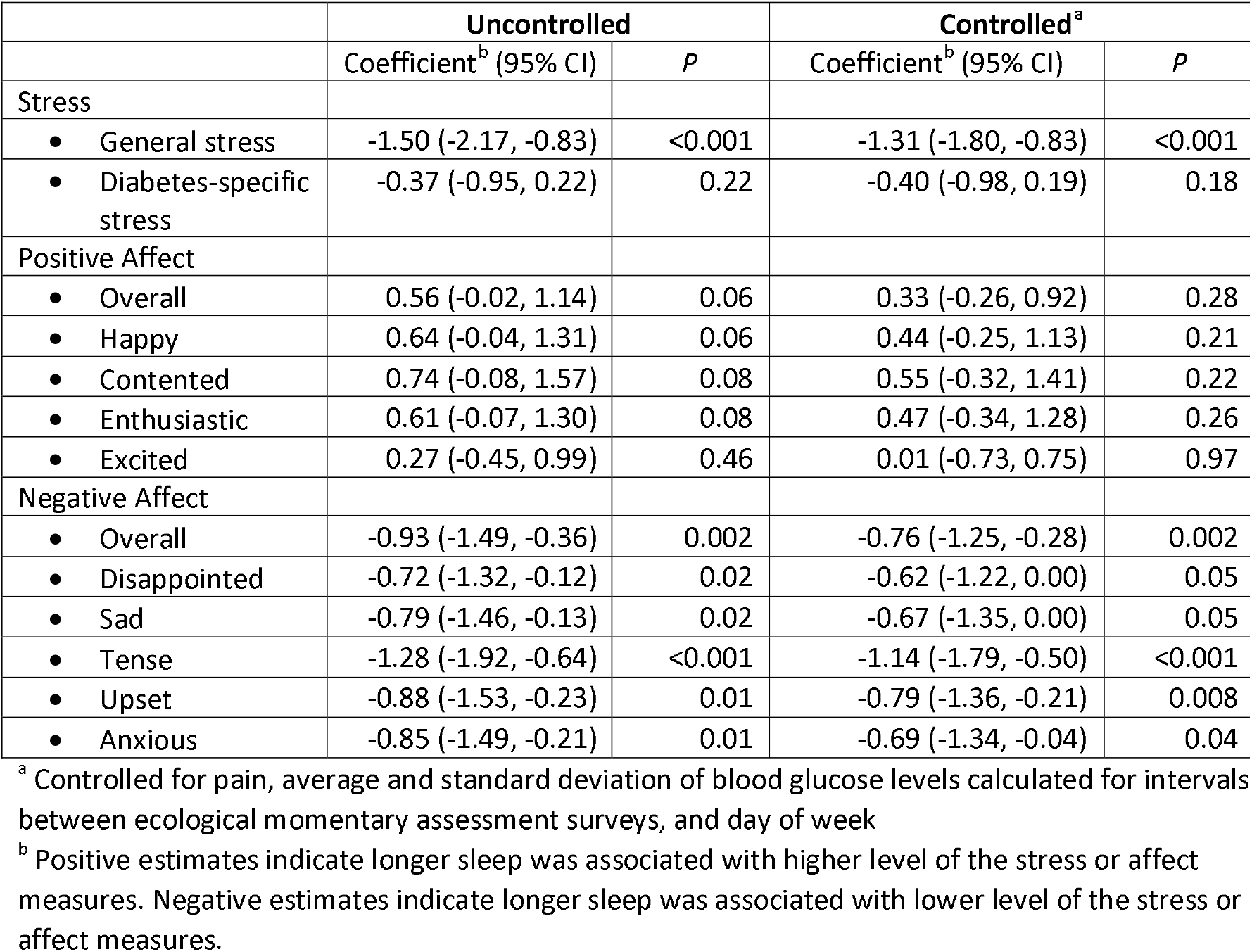
Within-person Relationship Between Sleep Duration and Next-Day Affect and Stress Among Individuals with Type-1 Diabetes (N=166)

|  | Uncontrolled |  | Controlled<sup>a</sup> |  |
| --- | --- | --- | --- | --- |
|  | Coefficient<sup>b</sup> (95% CI) | *P* | Coefficient<sup>b</sup> (95% CI) | *P* |
| Stress |  |  |  |  |
| • General stress | -1.50 (-2.17, -0.83) | <0.001 | -1.31 (-1.80, -0.83) | <0.001 |
| • Diabetes-specific stress | -0.37 (-0.95, 0.22) | 0.22 | -0.40 (-0.98, 0.19) | 0.18 |
| Positive Affect |  |  |  |  |
| • Overall | 0.56 (-0.02, 1.14) | 0.06 | 0.33 (-0.26, 0.92) | 0.28 |
| • Happy | 0.64 (-0.04, 1.31) | 0.06 | 0.44 (-0.25, 1.13) | 0.21 |
| • Contented | 0.74 (-0.08, 1.57) | 0.08 | 0.55 (-0.32, 1.41) | 0.22 |
| • Enthusiastic | 0.61 (-0.07, 1.30) | 0.08 | 0.47 (-0.34, 1.28) | 0.26 |
| • Excited | 0.27 (-0.45, 0.99) | 0.46 | 0.01 (-0.73, 0.75) | 0.97 |
| Negative Affect |  |  |  |  |
| • Overall | -0.93 (-1.49, -0.36) | 0.002 | -0.76 (-1.25, -0.28) | 0.002 |
| • Disappointed | -0.72 (-1.32, -0.12) | 0.02 | -0.62 (-1.22, 0.00) | 0.05 |
| • Sad | -0.79 (-1.46, -0.13) | 0.02 | -0.67 (-1.35, 0.00) | 0.05 |
| • Tense | -1.28 (-1.92, -0.64) | <0.001 | -1.14 (-1.79, -0.50) | <0.001 |
| • Upset | -0.88 (-1.53, -0.23) | 0.01 | -0.79 (-1.36, -0.21) | 0.008 |
| • Anxious | -0.85 (-1.49, -0.21) | 0.01 | -0.69 (-1.34, -0.04) | 0.04 |

<sup>a</sup> Controlled for pain, average and standard deviation of blood glucose levels calculated for intervals between ecological momentary assessment surveys, and day of week
<sup>b</sup> Positive estimates indicate longer sleep was associated with higher level of the stress or affect measures. Negative estimates indicate longer sleep was associated with lower level of the stress or affect measures.

**Table 3.** Estimates of Covariates in the Controlled Models with General Stress and Overall Negative Affect as the Dependent Variables

| Covariates | Model for General Stress |  | Model for Overall Negative Affect |  |
| --- | --- | --- | --- | --- |
|  | Coefficient (95% CI) | *P* | Coefficient (95% CI) | *P* |
| Pain | 5.09 (3.52, 5.10) | <0.001 | 3.88 (2.96, 3.99) | <0.001 |
| Blood glucose level, average | 0.30 (-0.10, 0.91) | 0.21 | 0.08 (-0.17, 0.48) | 0.62 |
| Blood glucose level, standard deviation | -0.04 (-0.38, 0.42) | 0.85 | 0.22 (-0.16, 0.36) | 0.09 |
| Day of week |  |  |  |  |
| • Monday | Reference |  | Reference |  |
| • Tuesday | 0.05 (-1.27, 1.37) | 0.94 | -0.17 (-1.38, 1.05) | 0.79 |
| • Wednesday | 1.48 (0.17, 2.79) | 0.03 | 0.80 (-0.44, 2.04) | 0.21 |
| • Thursday | 1.27 (-0.04, 2.58) | 0.06 | 0.58 (-0.67, 1.82) | 0.36 |
| • Friday | 1.15 (-0.15, 2.45) | 0.08 | 0.45 (-0.79, 1.69) | 0.47 |
| • Saturday | -1.76 (-3.08, -0.44) | 0.009 | -1.07 (-2.31, 0.18) | 0.09 |
| • Sunday | -3.00 (-4.32, -1.68) | <0.001 | -0.85 (-2.07, 0.36) | 0.17 |

The results of exploratory analyses for cross-level interactions of sleep duration with gender and baseline depression and anxiety are shown in Table 4. Examination of the interaction effects did not reveal significant differential relationships for men and women and for individuals with and without depression or anxiety at baseline.

**Table 4.** Cross-level Interactions of Sleep Duration with Gender and Baseline Depression and Anxiety

|  | Sleep Duration × Female |  | Sleep Duration × Depression at Baseline |  | Sleep Duration × Anxiety at Baseline |  |
| --- | --- | --- | --- | --- | --- | --- |
|  | Coefficient (95% CI) | *P* | Coefficient (95% CI) | *P* | Coefficient (95% CI) | *P* |
| Stress |  |  |  |  |  |  |
| • General stress | -0.94<br>(-2.32, 0.44) | 0.18 | 0.04<br>(-1.73, 1.83) | 0.96 | 0.27<br>(-1.84, 2.38) | 0.80 |
| Negative Affect |  |  |  |  |  |  |
| • Overall | -0.35<br>(-1.34, 0.64) | 0.49 | -0.41<br>(-1.68, 0.87) | 0.53 | -0.40<br>(-1.23, -0.20) | 0.61 |
| • Disappointed | -0.02<br>(-1.25, 1.22) | 0.98 | 0.00<br>(-1.59, 1.59) | 0.99 | -0.40<br>(-2.29, 1.48) | 0.68 |
| • Sad | -0.84<br>(-2.22, 0.54) | 0.23 | 0.12<br>(-1.70, 1.95) | 0.89 | 0.19<br>(-2.04, 2.41) | 0.87 |
| • Tense | -0.14<br>(-1.47, 1.18) | 0.83 | -0.23<br>(-1.93, 1.47) | 0.79 | -0.21<br>(-2.22, 1.81) | 0.84 |
| • Upset | -0.33<br>(-1.52, 0.86) | 0.59 | -0.58<br>(-2.10, 0.95) | 0.46 | -0.42<br>(-2.23, 1.39) | 0.65 |
| • Anxious | -0.55<br>(-1.88, 0.78) | 0.42 | -1.18<br>(-2.88, 0.53) | 0.18 | -1.03<br>(-3.06, 1.00) | 0.32 |

## 4. Discussion

To the best knowledge, this is the first study that examines within-person relationships between sleep duration and next-day stress and affect in the daily life of individuals with T1D. The results show that longer sleep is significantly associated with lower next-day general stress but not diabetes-specific stress, partially supporting our hypothesis. Findings from a recent study [16] indicate that higher self-reported sleep quality are significantly associated with reduced next-day general and diabetes-specific stress. The insignificant finding regarding diabetes-specific stress in this study may be caused by insufficiently large sample size; nevertheless, if there were a significant relationship as sample size becomes bigger, the current results suggest that the effect size of sleep on diabetes-specific stress might be smaller than general stress.

Our results demonstrate significant within-person relationships between sleep duration and next-day NA, including the overall NA level and all five NA items. The results reveal no significant within-person relationships between sleep duration and next-day PA. These findings partially support our hypothesis. As mentioned above, existing research based on healthy adults generally found that sleep is associated with both PA and NA, with the associations with PA being stronger than those with NA [21–24]. We speculate that the presence of T1D may possibly alter the relationships between sleep and affect, shifting the primary impact of insufficient sleep from decreasing PA in healthy adults to increasing NA in individuals with T1D. The reasons of such altered relationships remain unclear and require further investigation of potential pathways linking sleep and affect in T1D.

Results from the examination of cross-level interactions reveal no significant interaction effects of sleep duration with gender and baseline depression and anxiety. The findings suggest sufficient sleep is equally important for males and females and for T1D patients with and without common mental health conditions like depression and anxiety to manage their stress and NA.

Previous research has demonstrated that individuals with T1D have poorer sleep quality and shorter sleep duration than people without T1D [40], possibly due to differences in neuroendocrine sleep architecture [41], and that sleep disturbances are associated with elevated blood glucose levels, poorer self-management, and impairments in behavioural and cognitive function within this population [13]. Coupled with this study’s findings on stress and affect, there is a clear need for more effective interventions to support sleep duration and quality as a modifiable target to improve functioning among people with T1D. While some promising recent studies have been effective in enhancing sleep among children and adolescents with T1D [42,43], to our knowledge there remains a gap in the literature regarding sleep promotion for adults with T1D.

This study has both strengths and limitations. This study is based on intensive longitudinal data collected from the daily life of individuals with T1D, which improves ecological validity of the study findings compared to laboratory-based studies. The sample size (i.e., 166 individuals with 14 days of repeated measures) is relatively large compared to other studies adopting similar intensive longitudinal designs. The measure of sleep duration was based on a weighted average of both objective and subjective measures, which may improve measurement accuracy. The study is limited by its observational nature; therefore, the examined associations should not be interpreted as causal relationships. However, existing laboratory based studies have accumulated evidence supporting sleep loss as a causal factor of increased stress and poorer emotions [9,23]. Another limitation of the study is that the data supporting our findings were collected during the COVID-19 pandemic, which has been shown interfere with people’s sleep, stress, and mental health [44–47]. This may limit the generalizability of findings to pre- and post-pandemic times.

In conclusion, this study examined the within-person relationships between sleep duration and next-day stress and affect using intensive longitudinal data collected from the daily life of individuals with T1D. The study reveals that longer sleep was associated with lower next-day general stress and reduced NA. Results from the examination of cross-level interaction effects suggest longer sleep may equally benefit males and females and T1D patients with and without comorbid depression or anxiety. These findings suggest optimizing sleep duration as an important interventional target for better managing stress and improving daily emotional wellbeing of individuals with T1D.

## Data Availability

All data produced in the present study are available upon reasonable request to the authors.

## Acknowledgement

This research was supported by the US National Institute of Diabetes and Digestive and Kidney Diseases at the NIH (NIH/NIDDK #1R01DK121298-01).

