## Appendix Table for "Within-person Relationships of Sleep Duration with Next-Day Stress and Affect in the Daily Life of Adults with Type-1 Diabetes"

Appendix Table. Estimates of Covariates in the Models Examining the Within-Person Relationships between Sleep Duration With Next-Day Stress and Affect

| **Covariates** | **Model for General Stress** | | **Model for Diabetes-Specific Stress** | |
| --- | --- | --- | --- | --- |
|  | Coefficient (95% CI) | *P* | Coefficient (95% CI) | *P* |
| Pain | 5.09 (3.52, 5.10) | <0.001 | 3.15 (2.52, 3.79) | <0.001 |
| Blood glucose level, average | 0.30 (-0.10, 0.91) | 0.21 | 1.33 (0.94, 1.72) | <0.001 |
| Blood glucose level, standard deviation | -0.04 (-0.38, 0.42) | 0.85 | 0.66 (0.34, 0.98) | <0.001 |
| Day of week |  |  |  |  |
| - Monday | Reference |  | Reference |  |
| - Tuesday | 0.05 (-1.27, 1.37) | 0.94 | -0.27 -1.75, 1.22) | 0.73 |
| - Wednesday | 1.48 (0.17, 2.79) | 0.03 | 0.77 (-0.75, 2.28) | 0.32 |
| - Thursday | 1.27 (-0.04, 2.58) | 0.06 | 0.26 (-1.26, 1.77) | 0.74 |
| - Friday | 1.15 (-0.15, 2.45) | 0.08 | -0.46 (-1.97, 1.05) | 0.55 |
| - Saturday | -1.76 (-3.08, -0.44) | 0.009 | -1.48 (-3.00, 0.04) | 0.06 |
| - Sunday | -3.00 (-4.32, -1.68) | <0.001 | -1.63 (-3.12, -0.13) | 0.03 |
| **Covariates** | **Model for Overall Positive Affect** | | **Model for Happy** | |
|  | Coefficient (95% CI) | *P* | Coefficient (95% CI) | *P* |
| Pain | -2.57 (-3.15, -1.98) | <0.001 | -3.28 (-4.03, -2.53) | <0.001 |
| Blood glucose level, average | 0.06 (-0.29, 0.42) | 0.72 | 0.33 (-0.13, 0.79) | 0.16 |
| Blood glucose level, standard deviation | 0.36 (0.07, 0.65) | 0.01 | 0.31 (-0.07, 0.69) | 0.11 |
| Day of week |  |  |  |  |
| - Monday | Reference |  | Reference |  |
| - Tuesday | -0.58 (-2.05, 0.89) | 0.44 | 0.11 (-1.65, 1.86) | 0.91 |
| - Wednesday | 0.44 (-1.07, 1.95) | 0.57 | -0.30 (-2.09, 1.49) | 0.74 |
| - Thursday | -0.04 (-1.54, 1.47) | 0.96 | -0.33 (-2.12, 1.45) | 0.72 |
| - Friday | 1.29 (-0.22, 2.80) | 0.09 | 1.18 (-0.60, 2.97) | 0.19 |
| - Saturday | 2.89 (1.37, 4.40) | <0.001 | 3.10 (1.31, 4.90) | <0.001 |
| - Sunday | 1.55 (0.08, 3.02) | 0.04 | 2.06 (0.30, 3.82) | 0.02 |
| **Covariates** | **Model for Contented** | | **Model for Enthusiastic** | |
|  | Coefficient (95% CI) | *P* | Coefficient (95% CI) | *P* |
| Pain | -3.00 (-3.76, -2.23) | <0.001 | -2.33 (-3.12, -1.54) | <0.001 |
| Blood glucose level, average | -0.16 (0.63, 0.31) | 0.50 | -0.15 (-0.64, 0.34) | 0.54 |
| Blood glucose level, standard deviation | 0.21 (-0.17, 0.60) | 0.28 | 0.34 (-0.06, 0.74) | 0.10 |
| Day of week |  |  |  |  |
| - Monday | Reference |  | Reference |  |
| - Tuesday | -1.10 (-2.91, 0.71) | 0.24 | -0.56 (-2.37, 1.24) | 0.54 |
| - Wednesday | -0.91 (-2.75, 0.93) | 0.33 | 1.69 (-0.14, 3.52) | 0.07 |
| - Thursday | 0.09 (-1.75, 1.93) | 0.93 | 0.10 (-1.73, 1.93) | 0.91 |
| - Friday | -0.03 (-1.87, 1.81) | 0.97 | 1.97 (0.14, 3.80) | 0.03 |
| - Saturday | 1.95 (0.09, 3.80) | 0.04 | 3.18 (1.34, 5.03) | <0.001 |
| - Sunday | 0.97 (-0.85, 2.79) | 0.30 | 1.99 (0.18, 3.81) | 0.03 |

Appendix (Continued). Estimates of Covariates in the Models Examining the Within-Person Relationships between Sleep Duration With Next-Day Stress and Affect

| **Covariates** | **Model for Excited** | | **Model for Overall Negative Affect** | |
| --- | --- | --- | --- | --- |
|  | Coefficient (95% CI) | *P* | Coefficient (95% CI) | *P* |
| Pain | -1.84 (-2.65, -1.02) | <0.001 | 3.88 (2.96, 3.99) | <0.001 |
| Blood glucose level, average | 0.30 (-0.21, 0.80) | 0.25 | 0.08 (-0.17, 0.48) | 0.62 |
| Blood glucose level, standard deviation | 0.53 (0.12, 0.95) | 0.01 | 0.22 (-0.16, 0.36) | 0.09 |
| Day of week |  |  |  |  |
| - Monday | Reference |  | Reference |  |
| - Tuesday | -0.62 (-2.52, 1.27) | 0.52 | -0.17 (-1.38, 1.05) | 0.79 |
| - Wednesday | 1.30 (-0.63, 3.22) | 0.19 | 0.80 (-0.44, 2.04) | 0.21 |
| - Thursday | 0.14 (-1.78, 2.06) | 0.89 | 0.58 (-0.67, 1.82) | 0.36 |
| - Friday | 2.19 (0.27, 4.11) | 0.03 | 0.45 (-0.79, 1.69) | 0.47 |
| - Saturday | 3.50 (1.57, 5.44) | <0.001 | -1.07 (-2.31, 0.18) | 0.09 |
| - Sunday | 0.93 (-0.97, 2.83) | 0.34 | -0.85 (-2.07, 0.36) | 0.17 |
| **Covariates** | **Model for Disappointed** | | **Model for Sad** | |
|  | Coefficient (95% CI) | *P* | Coefficient (95% CI) | *P* |
| Pain | 4.50 (3.82, 5.17) | <0.001 | 3.39 (2.80, 3.99) | <0.001 |
| Blood glucose level, average | 0.33 (-0.08, 0.75) | 0.12 | -0.08 (-0.45, 0.29) | 0.67 |
| Blood glucose level, standard deviation | 0.03 (-0.31, 0.37) | 0.86 | 0.13 (-0.17, 0.42) | 0.41 |
| Day of week |  |  |  |  |
| - Monday | Reference |  | Reference |  |
| - Tuesday | -0.33 (-1.88, 1.22) | 0.67 | -0.37 (-1.85, 1.10) | 0.62 |
| - Wednesday | 1.01 (-0.57, 2.58) | 0.21 | 0.11 (-1.39, 1.62) | 0.88 |
| - Thursday | 1.03 (-0.54, 2.60) | 0.20 | 0.74 (-0.76, 2.25) | 0.33 |
| - Friday | 0.07 (-1.50, 1.63) | 0.94 | 0.18 (-1.32, 1.70) | 0.81 |
| - Saturday | -0.83 (-2.41, 0.75) | 0.30 | -0.55 (-2.06, 0.96) | 0.48 |
| - Sunday | -0.97 (-2.52, 0.59) | 0.22 | -0.21 (-1.69*, 1.27) | 0.78 |
| **Covariates** | **Model for Tense** | | **Model for Upset** | |
|  | Coefficient (95% CI) | *P* | Coefficient (95% CI) | *P* |
| Pain | 4.40 (3.67, 5.13) | <0.001 | 4.11 (3.44, 4.78) | <0.001 |
| Blood glucose level, average | 0.29 (-0.16, 0.74) | 0.21 | 0.19 (-0.22, 0.61) | 0.36 |
| Blood glucose level, standard deviation | 0.40 (0.03, 0.77) | 0.04 | 0.20 (-0.15, 0.54) | 0.26 |
| Day of week |  |  |  |  |
| - Monday | Reference |  | Reference |  |
| - Tuesday | 0.24 | 0.78 | -0.89 (-2.39, 0.61) | 0.25 |
| - Wednesday | 1.46 | 0.09 | 0.08 (-1.43, 1.60) | 0.91 |
| - Thursday | 1.23 | 0.15 | -0.80 (-2.32, 0.71) | 0.30 |
| - Friday | 1.15 | 0.18 | -0.25 (-1.76, 1.26) | 0.75 |
| - Saturday | -0.95 | 0.27 | -1.36 (-2.88, 0.16) | 0.08 |
| - Sunday | -0.95 | 0.26 | -0.85 (-2.35, 0.66) | 0.27 |

Appendix (Continued). Estimates of Covariates in the Models Examining the Within-Person Relationships between Sleep Duration With Next-Day Stress and Affect

| **Covariates** | **Model for Anxious** | |
| --- | --- | --- |
|  | Coefficient (95% CI) | *P* |
| Pain | 3.42 | <0.001 |
| Blood glucose level, average | -0.35 | 0.13 |
| Blood glucose level, standard deviation | 0.27 | 0.16 |
| Day of week |  |  |
| - Monday | Reference |  |
| - Tuesday | 0.54 | 0.53 |
| - Wednesday | 1.23 | 0.15 |
| - Thursday | 0.66 | 0.45 |
| - Friday | 0.78 | 0.37 |
| - Saturday | -1.75 | 0.04 |
| - Sunday | -1.21 | 0.16 |
